# Dietary Practice and Associated Factors among Pregnant Women in Nono Woreda west shoa, Oromia, Ethiopia

**DOI:** 10.1101/2020.11.27.20239624

**Authors:** Dereje Bayissa Demissie, Tesfaye Erena, Tufa Kolola

## Abstract

**Background:** Pregnancy is a unique and critical stage of life during which extensive anatomical, physiological, biochemical and several other related changes take place. The everyday vitality prerequisites for health women of typical weight who have a modestly dynamic way of life, increment during pregnancy and depend on the trimester of the baby. Maternal undernutrition is a serious developmental challenge contributing a large share to the global disease burden. It is a major reason for the increased risk of adverse pregnancy outcomes, poor infant survival, and elevated risks of chronic diseases at later stages of life. Ethiopia has an unacceptably high burden of malnutrition and its consequences, and yet little is known about the determinants and responses to undernutrition during pregnancy

**Objectives:** To assess the dietary practice and associated factors among pregnant women in Nono Woreda, West Shoa Zone, Ethiopia.

**Methods:** A community-based cross-sectional study design with both quantitative and qualitative data collection was conducted. Simple random sampling was used to select 378 pregnant women. The data were collected using interviewer-administered questionnaire of Afan Oromo version. Data were entered using Epi info version 7 and analyzed by using Statistical Package for the Social Sciences (SPSS) software for Windows version 21. Multiple logistic regressions were run to assess factors that were associated with the dependent variable at P<0.05 and to control the confounders.

**Result:** Finally, the result of the study has shown that 31% of the study participants had good dietary practices while the rest 69% of pregnant women had poor dietary practices. Concerning dietary knowledge about a balanced diet, 63.5% of the study participants had good dietary knowledge while 36.5% had poor dietary knowledge about a balanced diet. Marital status, breastfeeding, health-seeking behavior, food avoiding, and dietary knowledge were shown to have a significant association (P < 0.05) with dietary practices of pregnant women. The quantitative study revealed that marital status (AOR =95%CI, 7.983(1.387, 45.947, P<0.02).

**Conclusion:** A dietary practice of pregnant women in the study area was poor. Marital statuses, breastfeeding, health-seeking behavior, food avoiding and dietary knowledge of balance diet were independent predictors of women dietary practices. Therefore, Health professional, Zonal health office, regional Bureau and health planners would be better to increasing awareness to have good dietary practices those women married and supported by household head.

## INTRODUCTION

### Background

The dietary habits of pregnant women are important for the proper progression of pregnancy and the development and health of the fetus (1). Proper food and good nutrition are essential for survival, physical growth, mental development, performance and productivity, health and wellbeing. It is a unique and critical stage of life during which extensive anatomical, physiological, biochemical and several other related changes take place (2). Pregnancy is a critical phase in a woman’s life when the expecting mother needs optimal nutrients of superior qualities to support the developing fetus naturally, the urge to eat more is experienced by nearly all pregnant women. However, the nutrition requirement varies with respect to age, gender and during physiological changes such as pregnancy(3). Malnutrition prevents individuals and even the whole country from achieving full potential and is closely related to survival, poverty, and development (4). Malnutrition has been identified as the major underlying contributing factor for nearly half (45%) of all child and a fifth of maternal deaths. Maternal and child undernutrition, is the leading global developmental challenge affecting nearly half of the world’s population and responsible for the death of 3.5 million mothers and children annually (5). Malnutrition accounts for 7% of the global disease burden and contributes to an increased probability of poor pregnancy outcomes. It has been strongly linked to increased risk of adverse pregnancy outcomes, poor infant survival, and risk of chronic diseases at later stages of life (6). Maternal undernutrition is a serious developmental challenge contributing a large share to the global disease burden. It is a major reason for the increased risk of adverse pregnancy outcomes, poor infant survival, and elevated risks of chronic diseases at later stages of life. Ethiopia has an unacceptably high burden of malnutrition and its consequences, and yet little is known about the determinants and responses to undernutrition during pregnancy (8). For instance, exposure to nutrition information, attitude towards specific dietary habits and nutrition knowledge, attending antenatal care, maternal education and income were explored as predictors of maternal dietary practices (9). In addition, age at first marriage, meal frequency, educational status, occupation of the head of household, religion, maternal age, and marital status, were discovered as predictors of maternal nutritional status which in turn influence dietary practices of mothers (10).

### Statement of the problem

The time during pregnancy is essential that the diet provides the energy and nutrients required to keep the mother healthy, to prevent pregnancy-related diseases and to allow the fetus to grow and develop in favorable conditions. Maternal nutrition continues to gain interest in many parts of the world (11). During pregnancy, the body’s need increases by 13% energy, 54% protein, 50% vitamins and minerals (12). Malnutrition is a serious public health challenge which has been directly associated with increased mortality and morbidity rate especially in many parts of developing countries. According to the World Health Organization, 585,000 deaths resulting from pregnancy and childbirth-related complications occur globally with about 1,500 deaths recorded daily, however, most of these deaths occur in developing countries (13). Globally, the study was done in China, 35% of the prevalence of anemia among pregnant women in the third trimester and also 3.9% is the prevalence of LBW (14). Maternal nutritional status is considered to be an important factor that affects the successful completion of pregnancy. In extreme cases of chronic undernutrition, low energy intake during pregnancy was associated with low birth-weight. However, the effect of moderate malnutrition on fetal growth is not clear (15).

A study done in Nigeria, shows that exact an incidence rate of 10–40% has been reported in a rural community in the northern part malnutrition during pregnancy. Also, 75% of pregnant women from the western part of Nigeria were reported to have had inadequate dietary energy intake. Poor nutrition in pregnancy negatively affects the woman’s health and that of the unborn child. Other interacting factors, such as racial and genetic background, age, general health, educational status, past nutritional status of the mother, parity, multiple pregnancies, climate, socioeconomic conditions related to sanitation and infections, and the availability of health services make interpretation of the association between maternal nutrition and fetal development difficult(16). The definitive negative outcome of poor prenatal health and nutrition, as well as inadequate care during pregnancy and delivery, is reflected in the high prevalence of maternal mortality in developing countries; nearly 600,000 women die each year from pregnancy-related causes. Although poor prenatal nutrition contributes directly and indirectly to this large mortality rate, the extent of its contribution has not been measured because the main reported causes of maternal mortality (hemorrhage, obstructed delivery, eclampsia, sepsis, and unsafe abortion) greatly overshadow the role of nutrition itself (17). A study done in Gondar Town showed that good dietary practice was found to be 40.1% during pregnancy. Mothers education, monthly income, nutrition information, and dietary knowledge had a positive significant with pregnant mothers’ dietary practices (*P*<0.001). As dietary practices of pregnant mothers were relatively low in this study, the government in collaboration and strong integration with concerned bodies should be focused on providing nutritional education to increase the practices of pregnant mothers on maternal nutrition during pregnancy(18). This study is important in filling the gap in knowledge of dietary practice during pregnancy in the rural community of Ethiopia.

## Methods and materials

### Study Setting and Design

The study was conducted in West Shoa Zone’, Oromia Region, Ethiopia February 27to March 30, 2019. Nono woreda is one of the 22 Woreda which is found in West Shoa Zone, Oromia Regional state. The woreda has 33 rural kebeles and 2 urban kebeles.The total population of this woreda estimated to 116,334 female 56,193 and male is 60,141. The number of household estimated to 24,236. The number of women within childbearing age from 15-49 years is 21,673 of these the number of a pregnant mother is about 4,037. The woreda has the health service facilities, 4 health centers, and 33 functional health posts. A community-based cross-sectional study design with both quantitative and qualitative data collection was conducted among pregnant mothers.

### Sample Size Determination and Sampling procedure

#### The sample size was determined, using the prevalence of good dietary practice which is 33.9% (3)on maternal nutrition during pregnancy with 5% marginal error and 95% CI(confidence level) and a none response rate of 10 %. The required sample sizes became: 378

A cluster random sampling technique was used in sampling the study subjects. The District has 35 kebeles. Twelve kebeles were selected by simple random sampling from the total kebeles (the smallest sampling unit) all pregnant mothers are included in the sampling frame and out of which randomly selects using the random number proportional to the size of the community. The lists of pregnant women were accessed from respective health extension workers of selected kebeles. Once the sampling unit was listed, then simple random sampling by lottery method was implemented to identify the individual subject. The study subject would be all pregnant mothers who are selected in the study area. Pregnant women residing in the study area for at least six months prior to data collection and age greater than 18 years and above

#### Operational definitions and term

**Malnutrition** is a disease caused by not getting enough of the right food to eat or enough quantity of food. Causes of malnutrition are multi-factorial and can be divided into two immediately, underlying and basic.

**Antenatal care** is a type of preventive healthcare with the goal of providing regular check-ups that allow doctors or midwives to treat and prevent potential health problems throughout the course of the pregnancy while promoting healthy lifestyles that benefit both mother and child.

**Good Dietary practice** is dietary habit an individual to stay fit and well throughout her life. Good dietary mean having >=3 regular meals (i.e., breakfast, lunch, and dinner) which contain food groups/types, such as grains, fruits, vegetables, dairy products and protein foods. Good dietary for pregnant women also need to take a daily prenatal vitamin to obtain some of the nutrients that is hard to get from foods alone, such as folic acid and Iron.

**The poor dietary practice** is Poor maternal nutrition contributing to poor fetal development, which increases the risk that the baby born will be ill or die. During pregnancy the regular meals low (<3/day) and less food group/types consume, and micronutrient supplements and also energy, other nutrient intake does not increase, the body’s own reserves are used, leaving a pregnant woman (Nutrition Dietary Guide, 2010). According to this study, good dietary practice means that the scores of dietary practices were obtained by summation of each group of questions. The score of the respondents has taken and respondents were classified as having good dietary practice means core have >=3regular meals which represented by one (1) and poor dietary practices by taking their responses mean score have less than <3 regular meals which represented by zero (0)

Food Security is all people, at all times especially during pregnancy have physical and economic access to sufficient, safe and nutritious food that meets their dietary needs and food preferences for an active and healthy life food security construct from physical availability of food, economic and physical access to food, and adequate food utilization that is a function of the ability of the body to process and use nutrients as well as of the dietary quality and the safety of the foods consumed.

### Data collection procedure and Data Quality Assurance

A structured questionnaire was developed and adopted from EDHS 2011, food frequency questionnaire and WHO standard. Questionnaires were developed first in English then translated to Afan Oromo version. Data collectors were five trained female diploma nurses who can speak Afan Oromo and supervised by two trained BSC nurse /HO from the catchment HC. The questionnaires were pre-tested before the actual data collection is started. The data collection was conducted by the health professional from the existing health system to easily familiarize with the tool. The close follow-up by the very experienced supervisors was made whether the questionnaires was appropriately filled by the trained interviewers. Women were first to inform about the study and its aim, and those agreeing to participate was given written consent. A face-to-face interview was conducted with the participants.

To ensure the quality of data, two days training of data collectors and supervisors were undertaken and administration of the total sample size to assess its clarity, length, completeness, and consistency. The questionnaires were also translated into local language to facilitate understanding of the respondents.

The semi-structured questionnaires are pretested in health center unit. The pre-test is done on 5% of the total sample size. The questionnaire was then assessed for its clarity, length, and completeness. Some skip patterns are then corrected; questions difficult to ask were rephrased.

### Data analysis plan and management

Data was entered through using Statistical Package for Epi info version7 and then exported to SPSS Windows version 21.0for analysis. Then the data were cleaned and explored for outliers, missing values and any inconsistencies by visualizing, calculating frequencies and sorting. Corrections were made according to original data for further analysis descriptive, including. Finally, Analysis of univariate was done using frequencies & percent. Binary logistic regression and multivariable analysis were employed. Binary logistic regression analysis was used to examine the association between dependent and independent variables. All variables with p<0.05 in the binary logistic regression analysis were inserted into the multiple logistic regression model to identify factors independently associated with the dietary practice of pregnant women.

## Result

### Sociodemographic characteristics

A total of 378 pregnant women participated in this study making the response rate 99.7%. The mean age of study participants was 2.02 ± 0.6. About 62.7% of study participants were between the age ranges of 25–29. Concerning education status, 228(60.3%) was unable to read and write and 73% of study participants were housewives in occupational status.

Ninety-five percent of the participants were married. One hundred fifty-nine (42.1%) were orthodox by religion. Among the participants (66.1%) were Oromo by ethnicity. All most (47.9%) of the participants have a land size of less than one hectare. Among the respondents (59%) were family size less than 4.Majority (95.3%) of participants reported that household head decisions made by the husband. About 76.7%, 24.3% and 20.1% of participants have a radio, television and mobile phone, respectively. (Table 1)

**Table 1:**
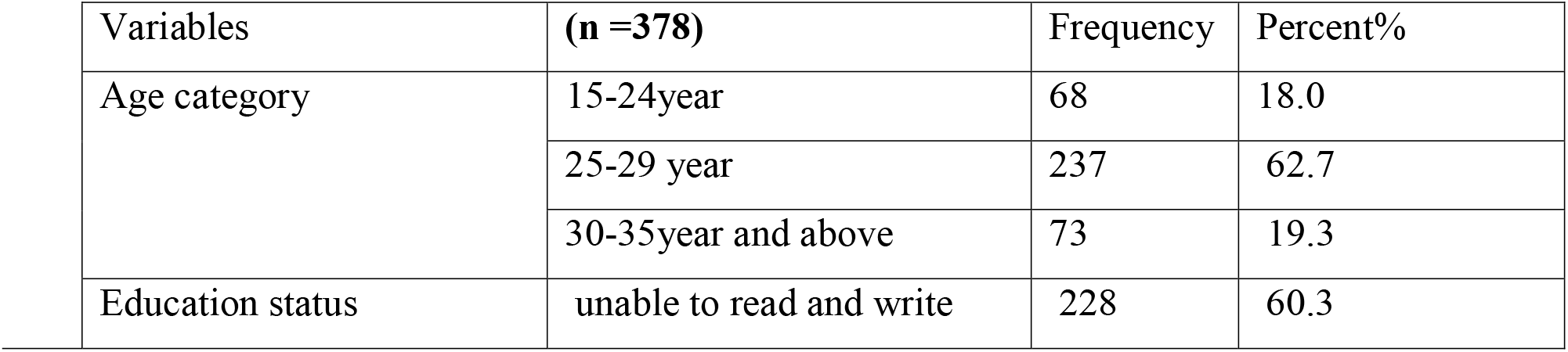
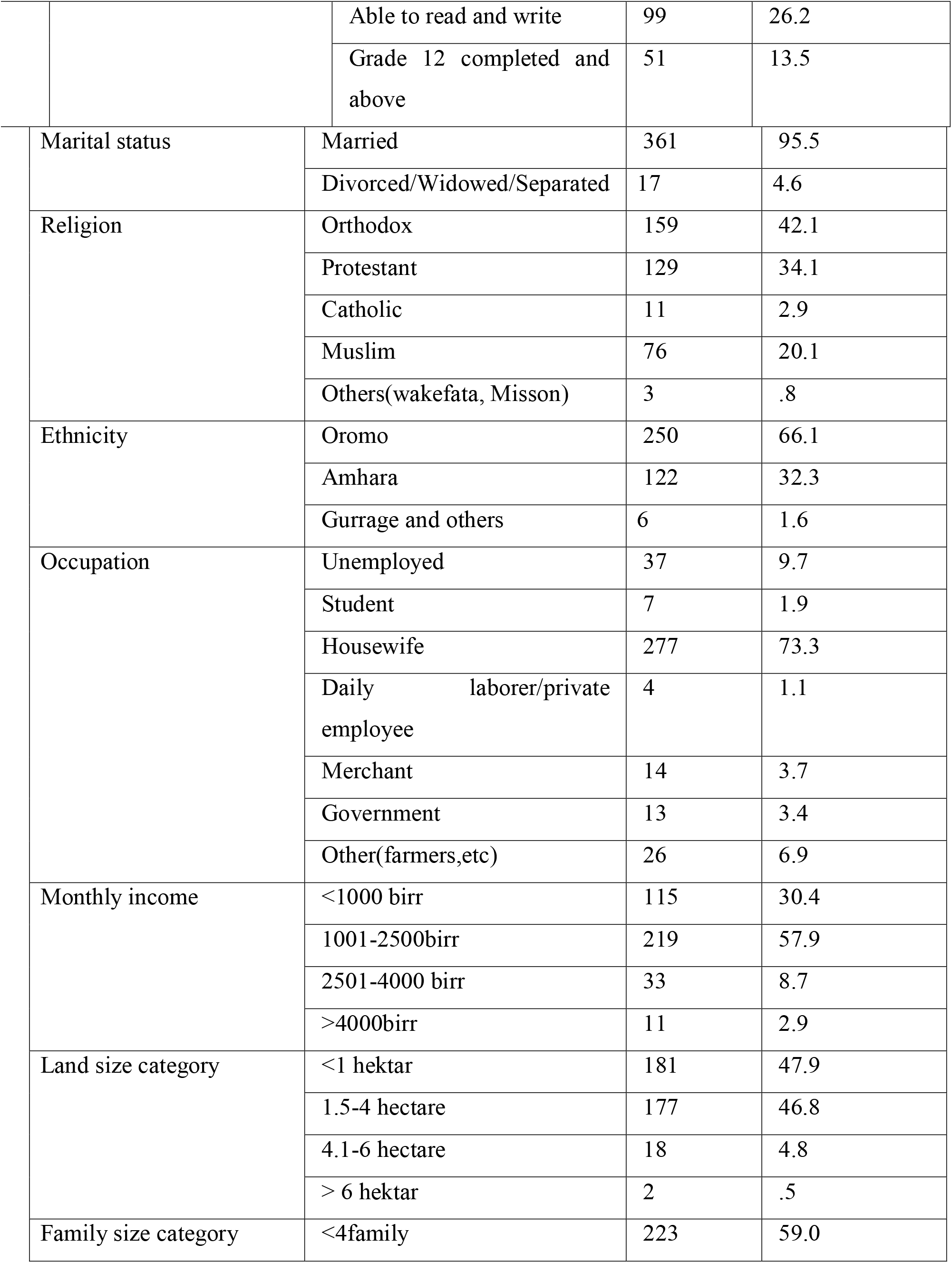
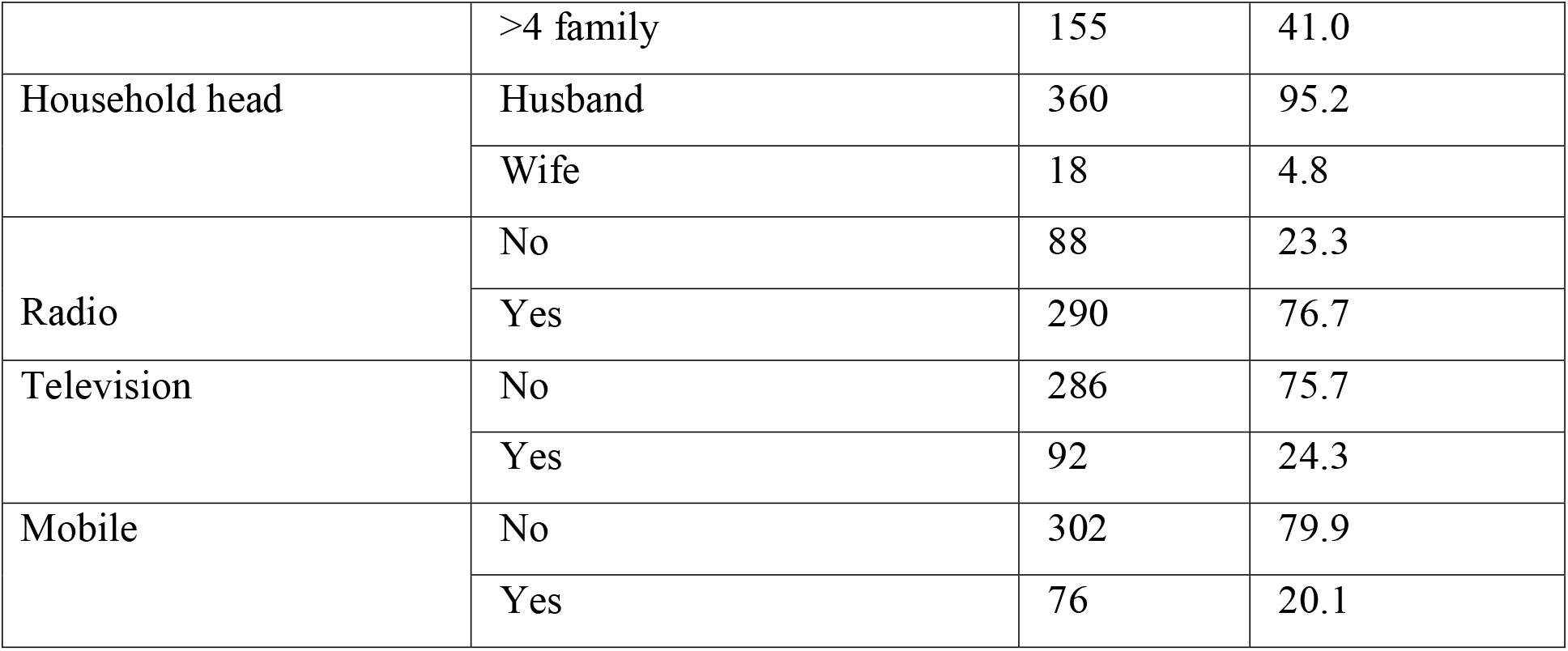
Socio- economic information characteristics of the pregnant women in Nono District, West Shoa Zone, Oromia Region State, Ethiopia,2019.

### Maternal and family characteristics of the study participants

During the study period, about 59.3% of study participants were gravid less than three.More than half of 194(51.3%) of the participants were between live birth range 1-4 children. Of total study participants, the majority 330(87.3%) had no history of abortion. From total study respondents 334 (88.4%) women had ANC (Antenatal care) follow up during current pregnancy; of which 325 (86%) had greater than or equal to three (3) times visit. Three hundred twenty-five (86%) of the participants given exclusive breastfeed their baby after birth. Among the respondents were 239(63.2%) safe water from the public tap and Three hundred thirty-six (88.9%) women were getting access to safe water in the area. The number of women who used medicine during pregnancy because of their preexisting condition and nausea/vomiting was 55 (14.6%) and 162(42.9%) respectively. There were 20(5.3%) and 2(0.5%) women who drank alcohol and smoked during current pregnancy respectively. One hundred sixty-one (42.6%) women were consuming coffee often (greater than or equal to two cups per day), 106(28%) seldom (once in a day) and61 (16.1%) none (don’t take coffee at all) (Table 2).

**Table 2:**
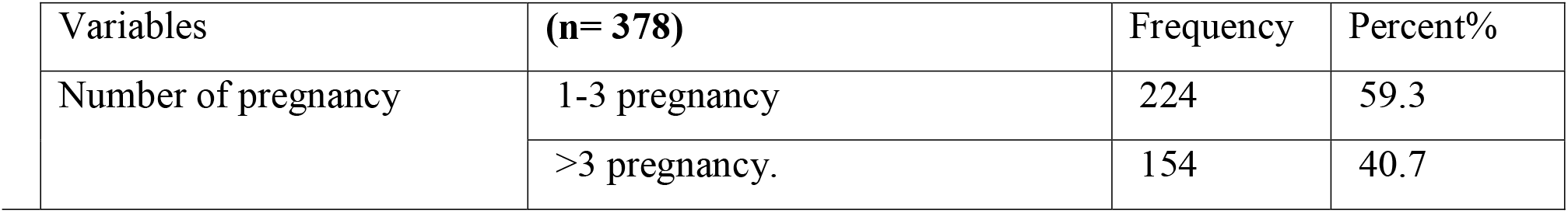
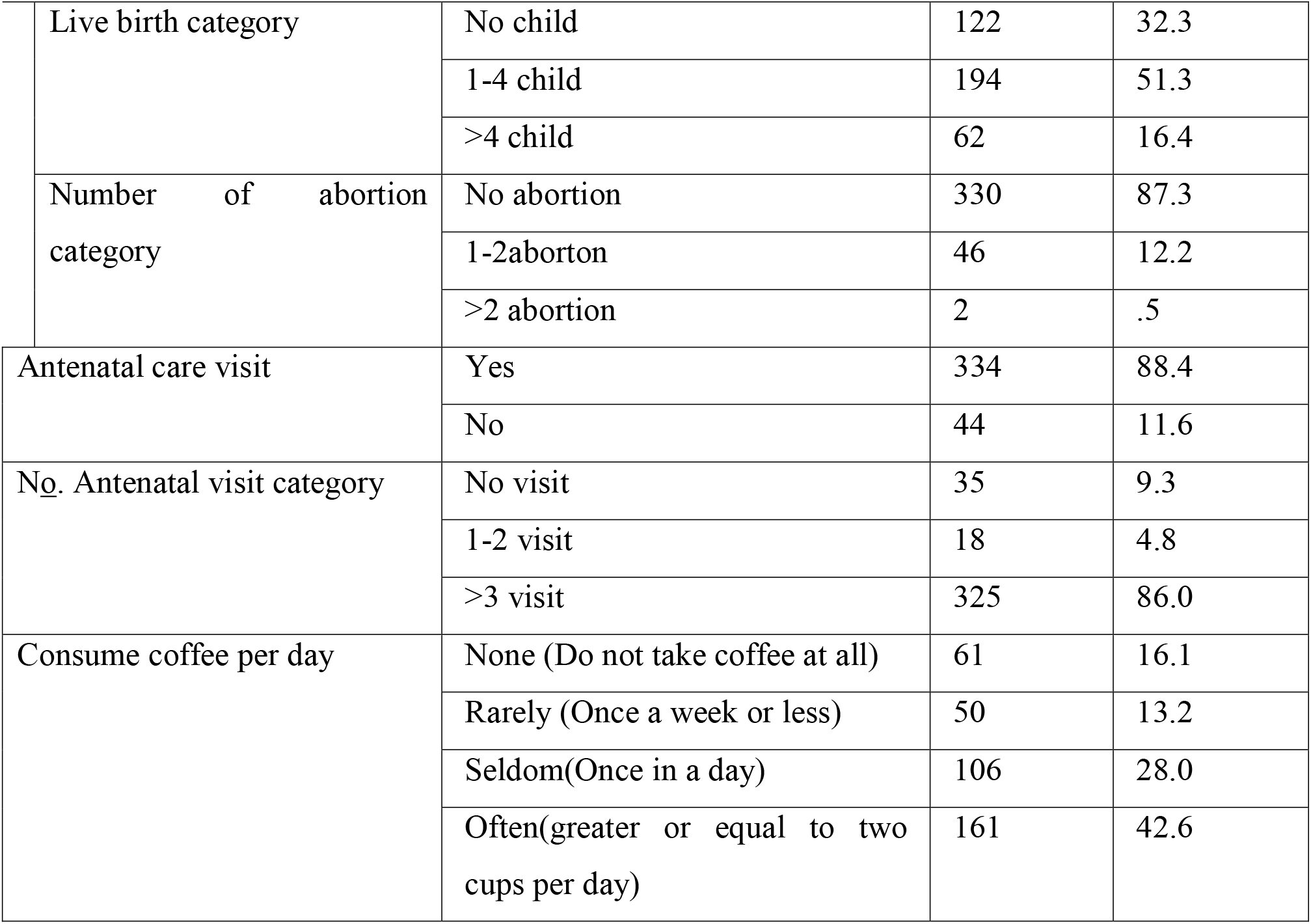
Obstetrics characteristics of the pregnant women in Nono District, West Shoa, Oromia Region, Ethiopia, 2019.

| Variables | (n= 378) | Frequency | Percent% |
| --- | --- | --- | --- |
| Number of pregnancy | 1-3 pregnancy | 224 | 59.3 |
|  | >3 pregnancy. | 154 | 40.7 |
| Live birth category | No child | 122 | 32.3 |
|  | 1-4 child | 194 | 51.3 |
|  | >4 child | 62 | 16.4 |
| Number of abortion category | No abortion | 330 | 87.3 |
|  | 1-2aborton | 46 | 12.2 |
|  | >2 abortion | 2 | .5 |
| Antenatal care visit | Yes | 334 | 88.4 |
|  | No | 44 | 11.6 |
| No. Antenatal visit category | No visit | 35 | 9.3 |
|  | 1-2 visit | 18 | 4.8 |
|  | >3 visit | 325 | 86.0 |
| Consume coffee per day | None (Do not take coffee at all) | 61 | 16.1 |
|  | Rarely (Once a week or less) | 50 | 13.2 |
|  | Seldom(Once in a day) | 106 | 28.0 |
|  | Often(greater or equal to two cups per day) | 161 | 42.6 |

**Table 3:** Dietary practices characteristics of the pregnant women in Nono District, West Shoa, Oromia Region, Ethiopia, 2019.

| Variables | (n = 378) | Frequency | Percent% |
| --- | --- | --- | --- |
| Change food in taking | Yes | 235 | 62.2 |
|  | No | 143 | 37.8 |
| Changed Food intake | Frequency of meals | 137 | 36.2 |
|  | Amount of meals per day | 125 | 33.1 |
|  | Both frequency and amount | 116 | 30.7 |
| Change the frequency of meals | Breakfast | 223 | 59.0 |
|  | Lunch | 100 | 26.5 |
|  | Dinner | 55 | 14.6 |
| Food group/types | Meat | 100 | 26.5 |
|  | Egg | 103 | 27.2 |
|  | Milk | 50 | 13.2 |
|  | Fruits | 83 | 22.0 |
|  | others,(vegetables etc) | 42 | 11.1 |
| Eating additional food | Yes | 185 | 48.9 |
|  | No | 193 | 51.1 |
| Avoiding eating any food | Yes | 257 | 68.0 |
|  | No | 121 | 32.0 |
| Food consuming per day cat | < 3per day | 198 | 52.4 |
|  | >=3 per day | 180 | 47.6 |
| Frequency of iron folate taking per day | Daily | 260 | 68.8 |
|  | Not daily | 78 | 20.6 |
|  | Not taking | 40 | 10.6 |
| Take Iron/folate | Yes | 288 | 76.2 |
|  | No | 82 | 21.7 |
|  | DK(do not know) | 8 | 2.1 |

**Figure 1:**
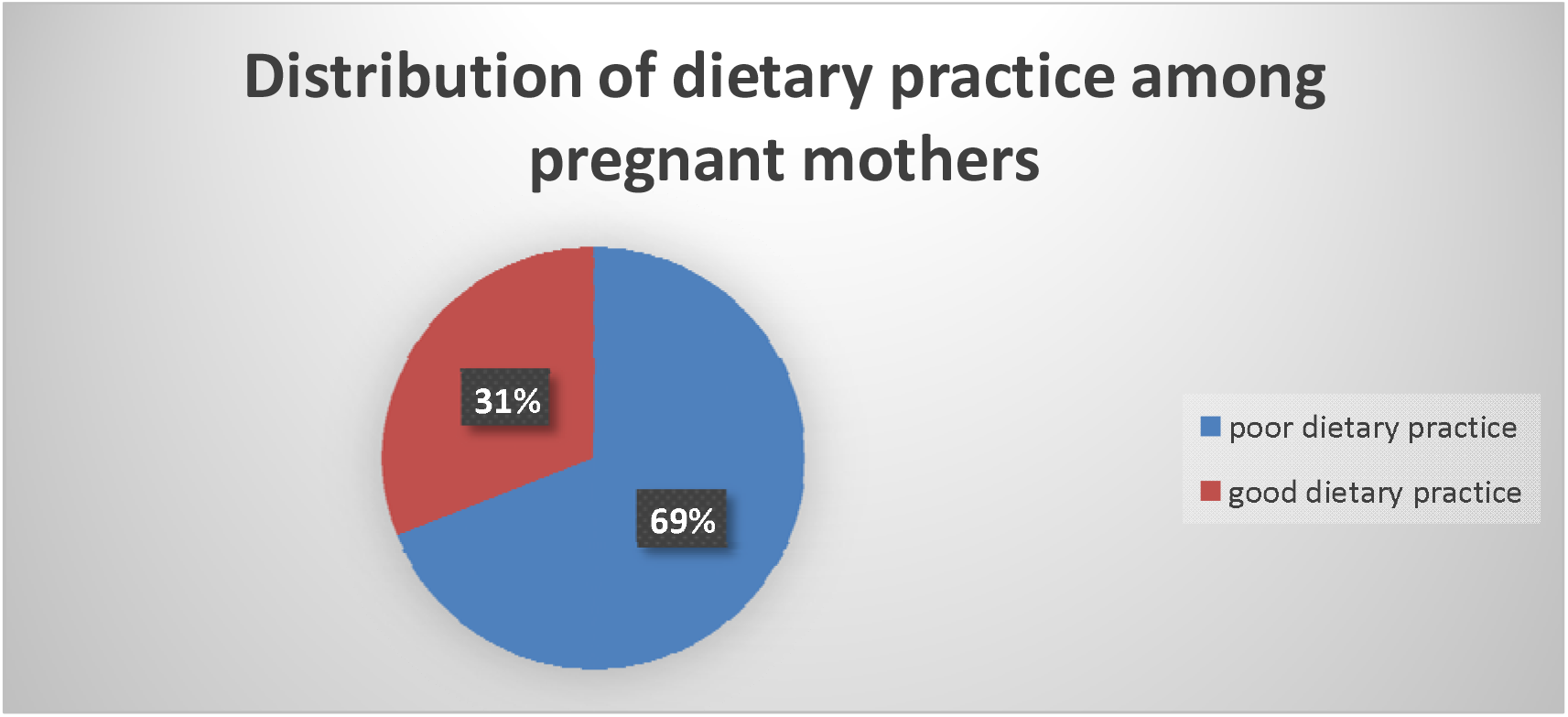
Distribution of dietary practice among pregnant mothers in Nono District, West Shoa, Oromia Region, Ethiopia, 2019.

### Dietary practices of the study participants

All most greater than half 235 (62.2%) women Changed food taking during the current pregnancy. From the participants, 137(36.2%) Changed the frequency of meals per day and 125(33.1%) amount of meals per day during pregnancy respectively. About 59% changed the frequency of meals their usual meal and the most commonly changed frequency of meals was breakfast. The result indicated that 257(68%) study participants avoid certain foods during pregnancy; of which103 (27.2%) egg, (26.5%) meat, (22%) fruits and (13.2%) milk avoids food during pregnancy. But (32%) of the respondents had not avoided any food item during their pregnancy. From study participants 185(48.9%) who took an additional meal during pregnancy. Concerning food consuming 198(52.4%) of the study participants were consuming less than 3 per day.

This study determined that the overall dietary practice of pregnant women revealed that the majority (69%) had poor dietary practices and 31% had good dietary practices (Figure 3).

### Dietary knowledge scores of pregnant women

The study showed that, 129(34.1%) women who were participants difficulty seeing dim light during pregnancy. In this study, 211(55.8%), 159(42.1%),84(22.2%)and72(19%) of the study participants had good dietary knowledge scores teff, vegetables, green vegetables, and milk, and also279(73.8%)teff,136(36%)greenvegetables50(13.2%)barley and47(12.4%)wheat for the knowledge about vitamin A source foods and iron source foods respectively, all other knowledge variables scored≥75% indicating good dietary knowledge. More than half 225(67.5%) of the study participants heard about goiter disease. Half of 189(50%), 112(29.6%)and 99(26.2(%)the study participants had known the major cause of goiter by not eating iodized salt, evil eye or evil spirit and drinking dirty water respectively.About123(32.5%),11329.9(%) and101(26.7%) had known the prevented method of goiter by doing tattooing(Nikisat), eating iodized salt and drinking holy water (tsebel). (Table 4)

**Table 4:**
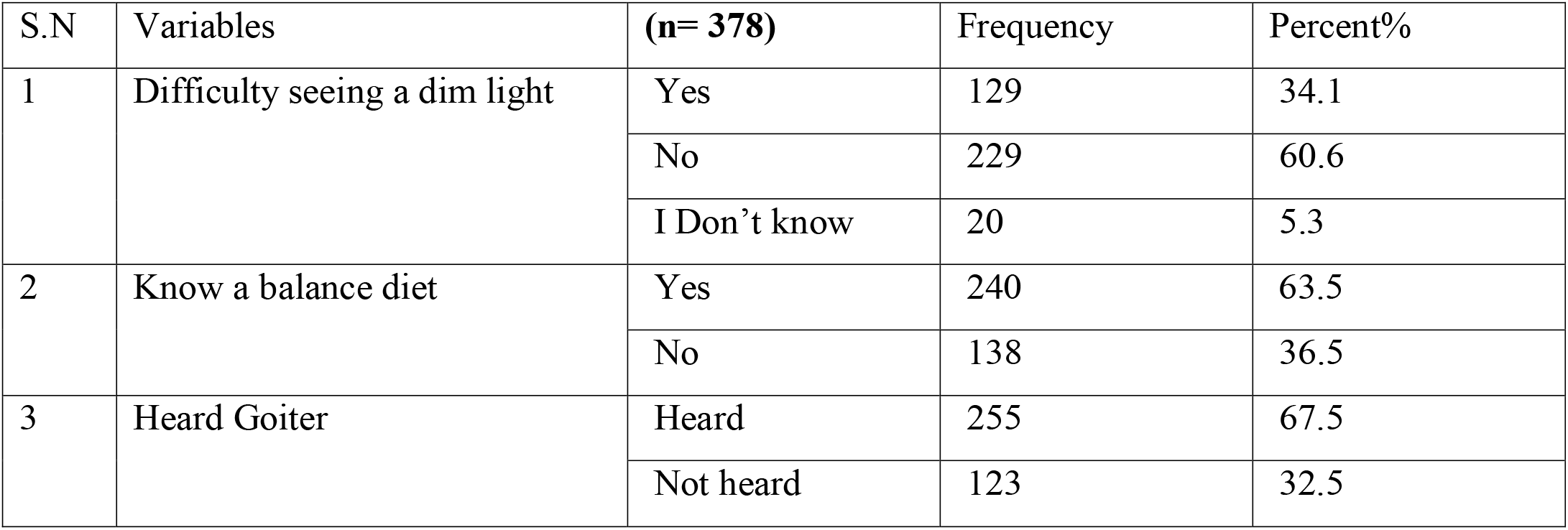
Dietary knowledge scores characteristics of the pregnant women in Nono District, West Shoa, Oromia Region, Ethiopia, 2019.

### Factors associated with dietary practices of pregnant women at multivariable logistic regression analysis

In multivariate logistic regression analysis, marital status, breastfeeding, health-seeking behavior, food avoiding and dietary knowledge during pregnancy of the women had a significant association with dietary practices (P < 0.05). Those study participants whose separated women were 8 times more likely to have good dietary practice than those married women.(AOR=7.983,95%CI,1.387,45.947).The study participants those who didn’t practice breastfeeding 2.8 times more likely to have good dietary practice than those practice breastfeeding(AOR=2.834, 95%CI, 1.426, 5.631). as well as women who didn’t health-seeking behavior 2.8 times more likely to have good dietary practice than those had health seeking behavior during pregnancy(AOR=2.800, 95%CI, 1.444, 5.41). The study participants who had food groups/types avoid fruits less likely to have good dietary practice than their counterparts during pregnancy (AOR=0.365, 95%CI, 0.150, 0.886). The study participants who have food balance diet knowledge were 4.7 times more likely to have good dietary practice than their counterparts.(AOR=4.739, 95%CI, 1.008, 2.999).(Table 5).

**Table 5:** Factors associated with dietary practices of pregnant women at multivariable logistic regression analysis in Nono District, West Shoa, Oromia Region, Ethiopia, 2019.

| Variables | Good dietary practice | Poor dietary practice | COR(95%,CI) | AOR | P-Value |
| --- | --- | --- | --- | --- | --- |
| Marital status |  |  |  |  |  |
| Married | 109 | 252 | 1.00 | 1.00 |  |
| Divorced | 3 | 6 | 1.156(0.284-4.706) | 0.607(0.112-3.303) |  |
| Widowed | 0 | 1 | (0.001) | 0.001(0.001-0.009) |  |
| Separated | 5 | 2 | 5.780(1.104 -30.251)* | 7.983(1.387-45.947)* | 0.020 |
| Previous Breastfeeding |  |  |  |  |  |
| Yes | 91 | 234 | 0.373(0.203-0.687) * | 1.00 |  |
| No | 25 | 24 | 1.00 | 2.834(1.426-5.631)* | 0.003 |
| Food group/types avoiding |  |  |  |  |  |
| Meat | 39 | 61 | 1 | 1.00 |  |
| Egg | 32 | 71 | 0.705(0.395-1.258) | 0.579(0.303-1.109) |  |
| Milk | 14 | 36 | 0.608(0.291-1.271) | 0.476(0.205-1.105) |  |
| Fruits | 20 | 63 | 0.497(0.261-.0945)* | 0.613(0.285-1.316) |  |
| Others,(vegetables etc) | 12 | 30 | 0.626(0.287-1.366) | 0.365(0.150-0.886)* | 0.026 |
| Health seeking behavior(Rx) |  |  |  |  |  |
| Yes | 18 | 67 | 1.00 | 1.00 |  |
| No | 99 | 194 | 1.899(1.070 -3.372) * | 2.800(1.444,5.41)* | 0.002 |
| Nutrition information/know balance diet |  |  |  |  |  |
| Yes | 57 | 81 | **1.00** | 1.00 |  |
| No | 60 | 180 | 2.111(1.350 -3.302) * | 4.739(1.008,2.999)* | 0.047 |

## Discussion

Dietary practices of pregnant women during pregnancy can determining the long-term health and nutritional status of both the mother and her growing fetus. This study determined that only 31% of the study participants had good dietary practices. This figure is lower than the study conducted in Bahir Dar town, Northwest Ethiopia,(28) but nearly similar with study in Guto Gida district, western Ethiopia(27). The differences may be due to Ethiopian government is currently promoting nutrition-related interventions through a health extension program, health facility nutrition services, community-based program and active involvement of pregnant women in focused antenatal care as well as in one-five network meeting at the community level.

In this study more than half of the study participants reported to taking no additional meals during pregnancy and lower prevalence of compared to study in Wondo Genet district, southern Ethiopia(30) the differences might be attributable to in sample size, geographical location, socio-economic characteristics, and cultural differences. This study showed that the diet frequency of meal per day: nearly half of the respondents (47.6 %%) had diet frequency of meal >=3 per day during their pregnancy. The rest (52.4%) had diet frequency of meals less than 3 per day during their pregnancy. The figure resulted by this study about frequency meal consumption of less than three per day is lower than the study conducted in Wondo Genet district, southern Ethiopia and Ghana, that a greater proportion (37.7%) of the women are more than three times during pregnancy as compared to only 11.5% before pregnancy (26). This study showed that (68%) of the respondents had practiced avoiding food during their pregnancy. Out of those who avoided food during their pregnancy, (27.2%) reported that it avoids the type of food they mentioned were an egg, (26.5%): type of food they mentioned was meat and, (22%) reported makes fruits the reason for avoidance of the food respectively. This figure is greater than the study conducted in Wondo Genet district, southern Ethiopia (30) .in which three fourth (75.7%) of the pregnant women in the study avoided at least one kind of food. This discrepancy may be the difference in the economic background of the study participants in the studies area. This might be due to the fact that the more husband earns, the more he invests on family nutrition and health which in turn attributes to good dietary practices of the family in general and pregnant women in particular. This study is in line with other studies which also reported that income has a positive significant association with women dietary practices.

## Conclusion

In this study only 31% of the study participants had good dietary practice indicating that the general dietary practice of the pregnant women in Nono District is not most desirable and the rest 69% of pregnant women reported poor dietary practices. Concerning dietary knowledge about balance diet, 63.5% of the study participants had good dietary knowledge while 36.5% had poor dietary knowledge about balance diet. Marital status, previous breastfeeding, health-seeking behavior, food avoiding and dietary knowledge were significantly associated with dietary practices of pregnant women in Nono District, West Shoa, Oromia Region, Ethiopia.

## Data Availability

All data underlying the findings described in this manuscript fully available without restriction and if the article is accepted for publication, the data availability statement will be published as part of the final article. There is no legal and/or ethical organization imposing restriction on this research article. Authors have no special access privileges to the data.
Data Sharing Statement
Data will be available upon request from the corresponding author

## Recommendations

Based on the findings of this research low proportion of good dietary practice during pregnancy to show, so how it would be improved or to give attention by the responsible bodies to identified the problem and how to improved knowledge towards dietary practice by health professional, Zonal health office, regional Bureau and health planners, like FMOH, NGO working this area and for researchers further study. Therefore, increasing awareness to have good dietary practices those women married and supported by household head.

Improving knowledge on balance diet through the government in with concerned bodies should focus on nutrition education, information about nutrition and integrating key nutrition messages nutrition during pregnancy in the study area

➢ Behavioral change communications that are culturally-adapted are needed to improving the intake of nutrient-dense foods as well as the compliance to supplements like Iron Foliate during pregnancy. An overall guideline that aims to improve the diets of pregnant women is needed.
➢ Behavioral change communications should aim to discourage the consumption of beverages like coffee during pregnancy to not further compromise the bioavailability of minerals.
➢ Combating harmful traditional practices, taboos, dietary restrictions and misconceptions related to the intake of adequate and quality foods during pregnancy is urgently needed. To this end, role modeling, community conversations, and other related activities could be used.
➢ An overall national dietary guideline, including pregnant women, should be developed to help the implementation of food-based strategies in the country.

## Acronyms and abbreviation

BMI: Body Mass Index.
EDHS: Ethiopian Demographic and health survey.
ETB: Ethiopian Birr
IUGR: Intra-Uterine Growth Restriction
LBW: Low birth weight.
NDA: Nutrition and Allergies
NE: Nutrition education
TDHS: Tanzania Demographic and Health Survey
SPSS: Statistical Package for the Social Sciences
UNICEF: United Nations International Children’s Emergency
WHO: World Health Organization.

## Acknowledgements

We want to ‘acknowledge Ambo University, College of medicine and health sciences for giving us this opportunity’. Our deepest gratitude goes to study participants, Nono Woreda Health office, supervisors and data collectors for their support. Last not list, I grateful for HEWS, Data collectors, supervisor, and West Shoa zone Health office for their valuable contribution and support.

## Author Contributions

All authors made substantial contributions to conception and design, acquisition of data, or analysis and interpretation of data; took part in drafting the article or revising it critically for important intellectual content; gave final approval of the version to be published; and agree to be accountable for all aspects of the work.

## Ethics and Consent Statement

‘Ethical clearance was obtained from the Ethical review committee of Ambo University CMHS (CMHS-R3012/2020). ‘Formal letter of cooperation’ was written to Nono District Health office and administration office to get permission to conduct the study in the woreda. Moreover, prior to commencing the study, a written informed consent was obtained from each respondent before data collection. In addition to this parental written consent was found from a parent of respondent. Confidentiality was maintained by omitting their name, and personal identification of participant was not compelled to the study. ‘The right to refuse was respected and information collected from this research project was kept confidential and the collected information was stored in a file, without the name of the study participant’.

## Human Ethics

The following information was supplied relating to ethical approvals (i.e., approving body and any reference numbers): All participants provided written informed consent, and that this study was conducted in accordance with the ‘Declaration of Helsinki. Consent for publication: Ambo University and Authors agreed for publication in a reputable journal’. (‘Agreement Ref number’: CMHS-R30112/2020).

## Data Sharing Statement

All data underlying the findings described in this manuscript fully available without restriction and if the article is accepted for publication, the data availability statement will be published as part of the final article. There is no legal and/or ethical organization imposing restriction on this research article. Authors have no special access privileges to the data.

### Data Sharing Statement

Data will be available upon request from the corresponding author

